# Passing the Test: A Model-based analysis of safe school-reopening strategies

**DOI:** 10.1101/2021.01.27.21250388

**Authors:** Alyssa Bilinski, Joshua A. Salomon, John Giardina, Andrea Ciaranello, Meagan C. Fitzpatrick

## Abstract

**Background:** The COVID-19 pandemic has induced historic educational disruptions. In December 2020, at least two-thirds of US public school students were not attending full-time in-person education. The Biden Administration has expressed that reopening schools is a priority.

**Objective:** To compare risks of SARS-COV-2 transmission in schools across different school-based prevention strategies and levels of community transmission.

**Design:** We developed an agent-based network model to simulate transmission in elementary and high school communities, including home, school, and inter-household interactions.

**Setting:** We parameterized school structure based on average US classrooms, with elementary schools of 638 students and high schools of 1,451 students. We varied daily community incidence from 1 to 100 cases per 100,000 population.

**Patients (or Participants):** We simulated students, faculty/staff, and adult household members.

**Interventions:** We evaluated isolation of symptomatic individuals, quarantine of an infected individual’s contacts, reduced class sizes, alternative schedules, staff vaccination, and weekly asymptomatic screening.

**Measurements:** We projected transmission among students, staff and families during one month following introduction of a single infection into a school. We also calculated the number of infections expected for a typical 8-week quarter, contingent on community incidence rate.

**Results:** School transmission risk varies according to student age and community incidence and is substantially reduced with effective, consistent mitigation measures. Nevertheless, when transmission occurs, it may be difficult to detect without regular, frequent testing due to the subclinical nature of most infections in children. Teacher vaccination can reduce transmission to staff, while asymptomatic screening both improves understanding of local circumstances and reduces transmission, facilitating five-day schedules at full classroom capacity.

**Limitations:** There is uncertainty about susceptibility and infectiousness of children and low precision regarding the effectiveness of specific prevention measures, particularly with emergence of new variants.

**Conclusion:** With controlled community transmission and moderate school-based prevention measures, elementary schools can open with few in-school transmissions, while high schools require more intensive mitigation. Asymptomatic screening should be a key component of school reopenings, allowing reopening at higher community incidence while still minimizing transmission risk.

## Introduction

COVID-19 prompted an unprecedented number of school closures during the spring of 2020. All 50 states recommended or mandated public school closures, impacting at least 124,000 US schools and over 55.1 million students (1). Reopenings have proven challenging, and as of December 9, 2020, only 33% of K-12 students were offered full-time in-person learning, a figure that has decreased since the fall (2). Where in-person schooling has been offered, a substantial percentage of families have opted out, including more than half in parts of Orange County, Florida and 70% in New York City (3,4). Nevertheless, many parents and advocates have objected strongly to school closures (5), and the Biden administration has stated a priority to rapidly facilitate safe, in-person school reopening (6).

Debates around school reopening have been heated and often invoked seemingly contradictory evidence about its safety. For example, a number of well-studied cases in school settings have found minimal secondary transmission (7–9). Nevertheless, significant school clusters have also been documented, particularly in Israel and in parts of the United States (10–12), and some observational studies have suggested that school closures may have substantially reduced transmission (13). Proponents of reopening schools point to evidence that COVID-19 is most often mild in children (15), while others have expressed concern that infection may lead to severe outcomes in some children, as well as among staff, families, and the community. The discord plays out similarly on a macro scale: a number of Asian and European countries reopened schools with physical distancing when community transmission was low and reported negligible increases in transmission (16,17). Some, including France, the United Kingdom, and Ireland, kept schools open during the fall wave and have reversed surging transmission by closing other sectors, although the latter two closed schools following the emergence of variant B.1.1.7 (18,19). Others, such as Austria, the Czech Republic, and South Korea, have closed schools to address rising case burdens (18).

Nevertheless, there is little debate that the benefits of in-person education are substantial, particularly amidst reports of high levels of remote absenteeism, increased depression, anxiety, and suicidality, and parent concerns around educational quality (20–22). Beyond educational and mental health outcomes, opening schools can also improve access to social services for children and labor market outcomes for working parents, especially women (23–27). In order to reopen safely, it is critical to take a comprehensive account of the full array of sometimes contradictory evidence and identify procedures to minimize risks.

We simulated SARS-CoV-2 transmission dynamics in elementary and high schools, to characterize how school transmission may occur either as isolated or sustained outbreaks. While several recent papers have discussed the impact of school reopening on the broader epidemic under different mitigation strategies (28–30), our work focused on the risk that transmission will occur on a school campus and spread to households of students and educators/staff. We emphasize uncertainty in transmission risk (rather than focusing on the average) as well as the observability of infections to school and public health staff, allowing us to reconcile apparent contradictions in the evidence. We evaluated outcomes under varying combinations of community incidence, in-classroom mitigation efforts, testing practices, and staff vaccination.

We project that consistently implemented mitigation, including masking and distancing, can drastically reduce the risk of school-based transmission, particularly in elementary schools. Nevertheless, high rates of subclinical disease, especially in children and adolescents, may render transmission difficult to connect to schools via contact tracing, and imperfect implementation of mitigation strategies or stochastic error (“bad luck”) can produce outbreaks even in well-managed systems. We find that teacher vaccination and asymptomatic screening can help support safe return to 5-day schooling for all children with only targeted closures, even amidst moderate ongoing community transmission.

## Methods

We developed an age-specific, agent-based network model of COVID-19 transmission in elementary and high school communities (Figure 1). We incorporated interactions within schools and homes, as well as those between households in childcare or social interactions outside of the school setting. Because data are particularly inconclusive on transmission among middle school-aged children, we did not model middle schools explicitly; accumulating evidence suggests that they may be more similar to high schools than elementary schools with respect to susceptibility and infectiousness (31,32).

**Figure 1:**
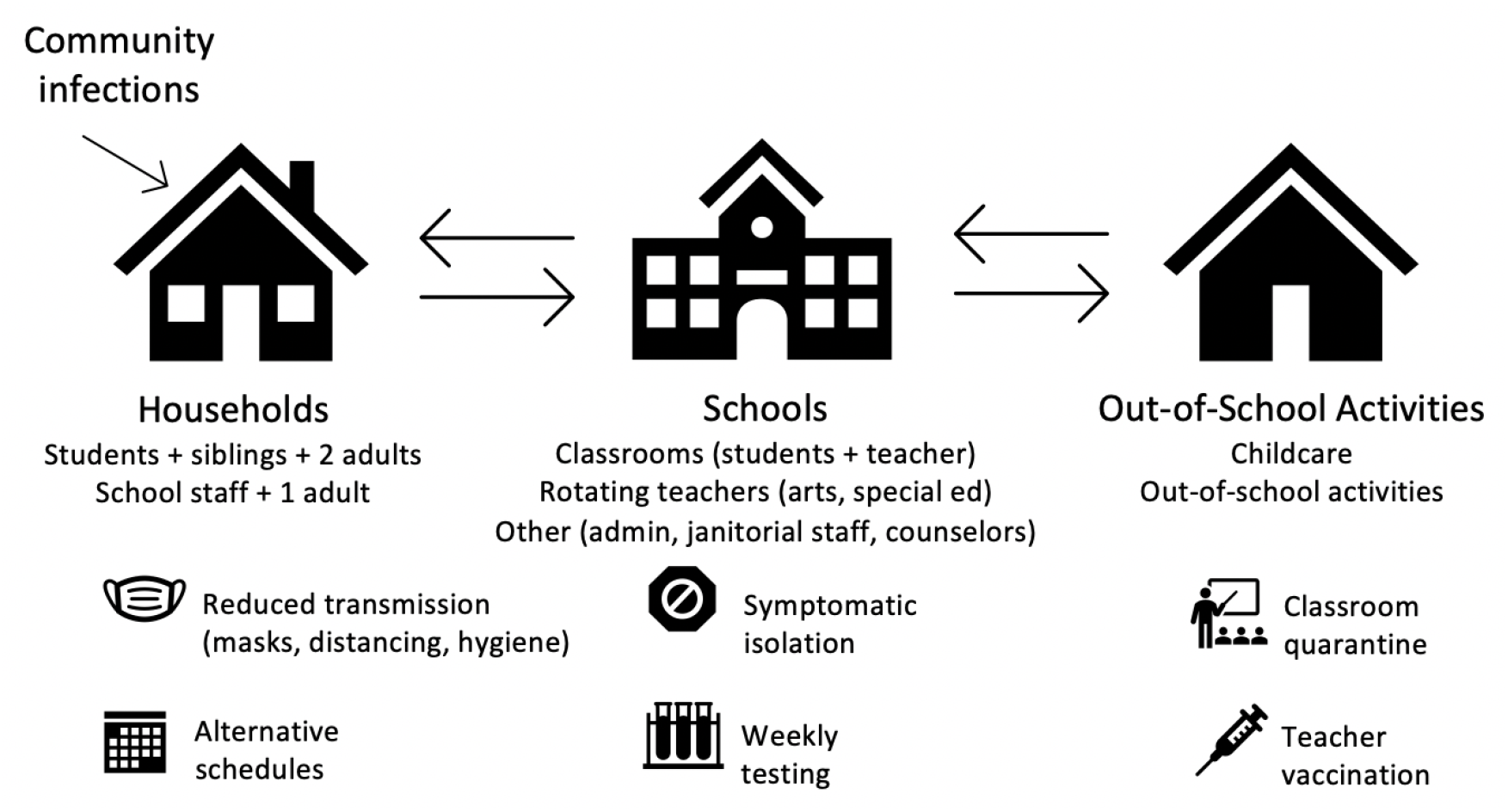
Model diagram. The model includes 3 primary domains: households, schools, and out-of-school social/childcare mixing and incorporates a range of interventions to prevent or reduce transmission.

### Scenarios

We simulated different combinations of interventions, incorporating strategies that target the three axes of general infection control in schools; COVID-19-specific countermeasures; and scheduling/cohorting with parameters detailed in Table 1.

**Table 1:** Parameter values.

| Parameter | Value | Source |
| --- | --- | --- |
| Number of students in elementary school | 638 | (37) |
| Average of students per elementary school class | 21.3 | Consistent with (37), 5 classes per grade |
| Number of adults who are not primary teachers in elementary schools | 30 | Estimate via school websites & personal communication |
| Number of students in high school | 1451 | (37) |
| Average of students per high school class | 23 | Consistent with (37), 16 classes per grade |
| Number of adults who are not primary teachers in elementary schools | 30 | Estimate via school websites & personal communication |
| Probability of asymptomatic disease for adults | 0.2 | (46) |
| Probability of asymptomatic disease for children | 0.4 | (8) |
| Relative infectiousness of asymptomatic disease | 0.5 | (46) |
| Probability of subclinical disease for adults (includes asymptomatic) | 0.4 |  |
| Probability of subclinical disease for children (includes asymptomatic) | 0.8 | (60) |
| Household attack rate | 20% (~4% per day) | (38) |
| Classroom attack rate | 1/2/3%<br>per day |  |
| Relative attack rate for random school contacts (vs. classroom) | 0.13 | Approximately 45 minutes |
| Relative attack rate for random staff contacts (vs. classroom) | 2 |  |
| Relative attack rate for childcare contacts (vs. classroom) | 2 | Assuming fewer precautions |
| Distribution on viral load | Lognormal<br>(.84, .3)/.84 | Produces 20%/45% from differences in viral load (47) |
| Incubation period (from exposure to symptoms, if symptomatic) | Gamma<br>(5.8, 0.95) | Estimated from (61) |
| Start of infectiousness relative to symptoms | Normal<br>(1.2, 0.4) | Estimated as mean 1.3 days, (63) mean 1.2 days w/lower bound of 95% CI of 3 for start and 2 for peak (64) |
| Duration of infectiousness | Lognormal<br>(5, 2) | (47) |
| Duration of isolation and quarantine | 10 days | (32) |
| Teacher vaccination uptake | 75% | (1,33) |
| Vaccination effectiveness | 90% | (35), assuming that vaccine blocks infections as well as prevents symptomatic illness (currently unknown) |
| Test uptake | 90% | Assumed |
| Test sensitivity during infectious period | 90% |  |

#### General infection control in schools

1. **Low uptake:** Schools implemented no or minimal general infection control measures, such as masking and distancing.
2. **Medium uptake:** Schools implemented masking and distancing, and adherence was moderate such that the risk transmission given infectious contact was 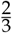 that experienced in a school with low uptake.
3. **High uptake:** Schools implemented masking and distancing and adherence was high, such that the risk of transmission given infectious contact was 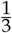 that experienced in a school with low uptake.

#### COVID-19-specific countermeasures

1. **Symptom-based self-isolation (“Symptomatic isolation”):** Individuals in the school were screened for symptoms daily, and those who developed clinically-recognizable symptoms did not attend school.
2. **Diagnostic testing plus classroom notification (“Classroom quarantine”):** Individuals who developed symptoms were isolated and immediately tested. The school received results within a day, and all classroom(s) associated with an individual who tested positive were notified and closed for 10 days, per CDC guidelines (32).
3. **Teacher vaccination with an infection-blocking vaccine (“Staff vaccination”):** In addition to classroom quarantine, teacher susceptibility was reduced to 33% of baseline, roughly assuming that 75% of teachers receive an infection-blocking vaccine with 90% effectiveness (33–35).
4. **Weekly asymptomatic screening (“Weekly screening”):** In addition to classroom quarantine, asymptomatic students and/or staff at each school were tested weekly. We assumed test acceptance was 90%, sensitivity was 90% during the infectious period, and results were available within 24 hours. Upon receipt of a positive result, infected individuals isolated outside of school for 10 days based on CDC guidelines, and siblings and classroom members of an individual who tests positive were notified and quarantined for 10 days.

#### Scheduling/Cohorting

1. **5-day schedule:** This scenario simulated a traditional 5-day, in-person learning schedule, allowing for classroom contacts, staff-staff interactions (10 per day), and random contacts between school members (20 per day). Related arts and special education teachers had revolving contact with 5 classrooms per day.
2. **Cohorting:** This scenario again assumed 5 days of in-person learning for all students, but with restricted out-of-classroom contacts, including separation of classes for lunch and recess. Students and primary teachers continued to have sustained classroom contacts. We assumed a 50% decrease in the number out-of-classroom contacts during the school day and that related arts were taught remotely.
3. **Half class sizes:** All students attended school five days per week, but in classes of half their typical size. To accommodate this, the number of teachers was doubled. Limited contacts as in (2) were also maintained.
4. **Hybrid scheduling (A/B):** Classes were subdivided into 2 cohorts, and students attended school for 2 days per week, either Monday/Tuesday or Thursday/Friday. Elementary school children in the same household attending the same school were sorted into the same cohort. All staff were physically present each instructional day. In sensitivity analysis, we considered alternative hybrid schedules. Limited contacts as in (2) were also maintained.

## Model structure

### Households

We generated a set of synthetic households from US population data (36) (Supplement). For elementary schools, we sampled from households containing at least one child aged 5 to 10 to identify siblings attending the same school. For high schools, we sampled from households with students aged 14 to 17. For each student, we included two adults in the household, based on the average number of household members over 25. For each staff member, we also included a household adult contact, representing a partner or roommate with whom they had close contact.

### Elementary schools

To simulate a representative school, we identified the average school size weighted by student population (37). We simulated an average-sized elementary school of 638 pupils from 500 households, of which 114 households had more than one child in the school. We set five classes per grade level, an average class size of 21 children, and one primary teacher per class. We also incorporated 30 additional staff, to reflect roles such as administrators, counselors, cafeteria staff, custodians, special education teachers, and teachers of related arts (or ‘specials’, e.g. music and art). These adults were assigned either in-classroom roles (n = 15, including special education and related arts teachers who are in rotating contact with entire classes of students) or out-of-classroom roles (n = 15, interacting with fewer individuals outside the classroom setting).

### High schools

We simulated an average-sized high school of 1451 students in 1225 households, with 142 households having more than one student in the school (37). Students rotated among 8 class periods per day (23 students and 1 teacher), with the distribution of students to class periods chosen randomly within grade levels but repeated daily for the full simulation. The high school had 45 additional staff with out-of-classroom roles and 15 with in-classroom roles. In both elementary and high schools, teachers were modeled to have additional staff contacts per day (e.g., contacts in break rooms or offices).

### Out-of-school interactions

Reflecting social interactions and out-of-school childcare, our base case analysis assumed that each family interacted with 1 additional family on each day they did not attend school, randomly reassorted each day. We varied this number in sensitivity analysis from 0 to 9 families per day out of school. We assume that when families mix, no more than 2 adults attend at a given time, who are randomly selected.

## Transmission and epidemiological parameters

At each daily time-step, we modeled dyadic interactions between individuals according to household, classroom, school, and childcare relationships, drawing parameter values from the distributions specified in Table 1. A SARS-CoV-2-infected individual i transmitted to susceptible individual j at time t with Bernoulli probability equal to:

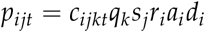

where *c*_*ijk*_ was an indicator variable equal to 1 if individuals *i* and *j* had contact type *k* at time *t, q*_*k*_ was the probability of transmission given one day of contact type *k, r*_*i*_ was the relative infectiousness of individual *i* (compared to full adult infectiousness), *s*_*j*_ was the relative susceptibility of individual *j* (compared to full adult infectiousness), and *a*_*i*_ indicated whether individual *i* had asymptomatic disease and *d*_*i*_ was a dispersion factor representing individual-level heterogeneity in transmissibility.

### Secondary attack rate (SAR) (q_k_)

Secondary attack rates (SARs) for SARS-CoV-2, the probability that a person with SARS-CoV-2 transmitted it to an individual they contacted, vary by contact type: household, classroom, random school, and out-of-school social/childcare contacts. Household SARs have been most frequently estimated in a number of contact tracing studies. While there is substantial variation across geographic locations, likely corresponding to cultural norms and precautions adopted, we assumed a 20% household adult-adult secondary attack rate over the full duration of an infection, translating to approximately 4% per day (38,39). This assumed minimal in-home quarantine measures but less household mixing than during full shelter-in-place orders.

We used the SAR from household contact tracing studies as a rough proxy for the per-day adult-adult attack rate in non-household settings, following close sustained contact in the absence of interventions such as masks and distancing. For school based transmission, we allowed adult-adult attack rates to reach up to 3% per day for scenarios with low uptake of masks and distancing, a downward adjustment from the household attack rate to account for less close contact in professional settings.^5^ We developed the scale of reductions in transmission from mitigation measures in line with observations from in household settings with high prevention measures (31,40) and based on effectiveness analyses for measures like masks (41,42).

### Relative susceptibility (s_i_) and infectiousness in children (r_i_)

Household contact tracing studies suggest that adults are likely more susceptible to COVID-19 and more apt to transmit it when infected than children, although data, particularly for the latter, are equivocal and limited by availability and timing of testing (43). Several sources indicate that any differences wane by the teenage years, possibly as early as age 10 (8,10,31,44,45). We assumed that elementary school children were half as susceptible as adults while high school children were equally susceptible. We likewise specified that elementary school children were half as infectious as adults and high school students were equally as infectious as adults (31,44). We discuss sources informing these assumptions further in the Supplement and varied them in sensitivity analyses.

### Asymptomatic transmission (a_i_) and overdispersion (d_i_)

We assumed that individuals with fully asymptomatic disease transmit COVID-19 at half the rate of those with any symptoms (46). While reported heterogeneity in transmission may be partially driven by differences in contact rates, we sample individual transmissibility according to a lognormal distribution to allow for some variation by individual characteristics (e.g. viral load) in adults who have been more widely identified as potential superspreaders (47,48).

## Model implementation

We first evaluated the downstream impact of a single infectious individual. We randomly designated one member of the school community as infected, starting on a random day of the week. We assessed the spread of the virus over 30 days, as in most cases either all infections were resolved over that time horizon or the spread was sufficiently large that additional public health measures (e.g. school closures) would be expected to be adopted during that time. For each scenario, we ran our model 2000 times and summarized:

1. Mean number of infections generated in the school over 30 days after index case
2. Percentage of scenarios without transmission from the index case
3. Percentage of scenarios with more than 5 in-school transmissions

We also describe the composition of secondary cases (proportion occurring in students, staff, and family members of students or staff).

Second, we quantified SARS-CoV-2 infections among our school population across a typical school quarter, given a constant community incidence. On each day over 8 weeks, every susceptible individual has a probability of becoming spontaneously infected outside of school that is equivalent to an age-adjusted community per capita daily incidence, distinct from their contact-dependent risk within the school network. When schools are remote, we assumed, similar to the analysis of hybrid schools, that each family interacted with 1 additional family on each day they did not attend school. For each scenario, we summarized:

1. Cumulative incidence overall and by type
2. Incremental incidence compared to remote learning

### Transmission control threshold

We defined a reopening strategy as controlling transmission in a group (students, educators/staff, or families) if it resulted in less than a one point increase in the percentage of the group which became infected, compared to remote learning. This threshold is broadly consistent with thresholds used by other modelers and with the consensus objective of minimizing in-school transmission (30,49).

## Results

### In-school transmission after the introduction of a single infection

#### In-school mitigation

In elementary schools with low mitigation and symptomatic isolation under a 5-day schedule, we projected an average of 2 secondary cases over 30 days following infection of a single index case (Figure 2A, left panel). This fell to 1 case with medium mitigation, and further to 0.4 cases with high mitigation. Under symptomatic isolation, transmission was most reduced by replacing the 5-day schedule with an A/B schedule (to 0.1-0.4 secondary cases depending on uptake of in-school prevention). With high mitigation, there were on average <0.5 secondary transmissions per case in all scenarios.

**Figure 2:**
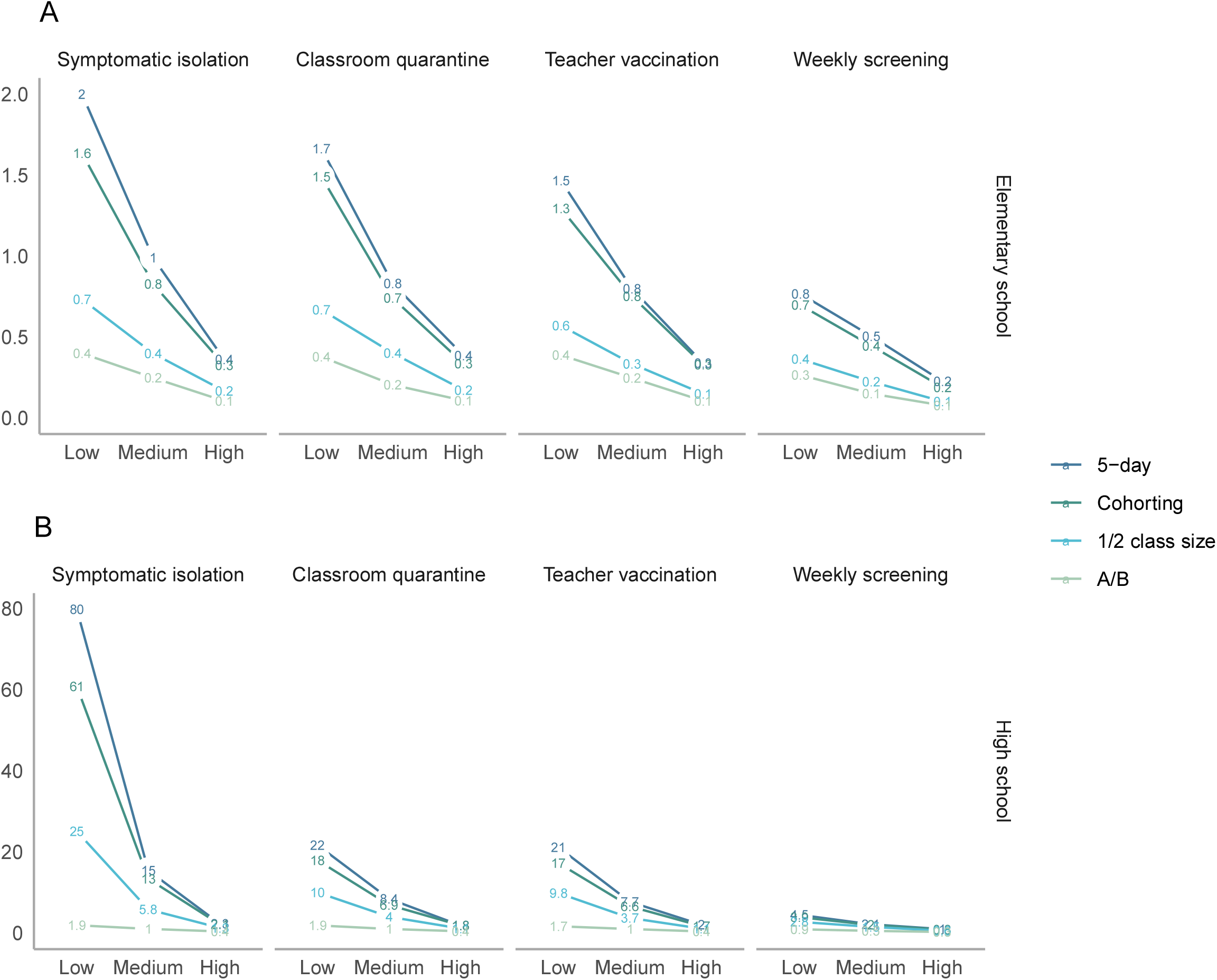
Average number of total secondary transmissions over 30 days (outside of the index case’s household) following a single introduction into a school community. (This is not an estimate of R, the effective reproduction number, which is displayed in Figure S8.) These include both transmission directly from the index case, as well as from secondary and tertiary cases. The top panel shows elementary schools, where children are assumed to be less susceptible and less infectious, while the bottom panel shows high schools. Note that axes differ across rows. The x-axes vary the level of prevention measure uptake, with low uptake assuming minimal interventions and high uptake assuming intensive interventions. Line colors correspond to scheduling strategies.

In high schools, where we specified that students acquired and transmitted infection at the same rate as adults, we found the potential for larger outbreaks after a single introduction into the school, particularly when uptake of in-school prevention was low (Figure 2B). For example, with low mitigation and classroom quarantine under a 5-day schedule, we projected 22 secondary cases in the school community over a 30-day period (in the absence of public health responses that would likely ensue before 30 days, such as school closure). High uptake of in-school mitigation reduced this markedly, to 1.8 cases.

### Quarantine, teacher vaccination, and screening

In elementary schools, where children were assumed to be less susceptible and infectious, classroom quarantine had a modest impact on transmission, although its impact was greater if the index case was a teacher (Figure S1). In high schools, classroom quarantine reduced projected average transmissions by a factor of 0.23 under low mitigation, but had smaller impact with high mitigation. We found a low impact of teacher vaccination on overall transmission, but substantial impact on transmission to teachers. Teacher vaccination reduced secondary infections over 30 days to 95% overall and 96% of the average without vaccination, in elementary and high schools respectively. In both settings, it reduced staff secondary incidence to a third of its initial rate, although the reduction was slightly greater if the index case was a teacher.

Weekly screening was projected to reduce secondary cases by a large degree compared to symptomatic isolation, classroom quarantine, or teacher vaccination: to an average of 0.2-0.8 with a 5-day schedule in elementary schools (varying by uptake of in-school prevention) and to 1.0-4.5 in high schools (Figure 2A, right panel). The impact of weekly screening following a single introduced case was greatest for settings with low mitigation: weekly screening reduced average projected secondary cases from 2.0 to 0.8 in elementary schools and from 80 to 4.5 in high schools.

### Detection of school-related transmission

Across all scenarios, for elementary schools, we estimated that 73% of secondary cases would occur in students, 19% in families, and 8% in teachers and staff. For high schools, 77% would occur in students, 19% in families, and 4% in teachers and staff (Figures S2 and S3). Although staff are the smallest of these percentages, because there are few staff relative to students and families, they had the highest per capita incidence of these groups. As children are less likely than adults to have symptoms, we projected that 14% of all secondary infections in elementary school communities and 15% in high school communities would be clinically symptomatic and therefore detectable without asymptomatic testing (Figure S4). With a more expansive definition of any symptoms, these percentages increased to 28% and 29% respectively. We discuss comparisons to documented in-school transmissions in the Supplement.

### Stochastic variation in secondary transmission

Re-running the simulation 2000 times with each set of parameters, we observed considerable variability across possible outcomes (Figure 3). In elementary schools with high mitigation and symptomatic isolation under a 5-day schedule, there was a 79% probability of zero secondary cases over a 30-day period (Figure 3A, left panel). However, there was a 0.3% probability of five or more secondary cases. The chance of >5 secondary cases occurring was higher when mitigation uptake was lower (Figure 3A, left and middle panels), reaching a 50% probability of zero secondary cases, and an 11% probability of five or more under low mitigation. In high schools with symptomatic isolation, the probability of no secondary cases similarly ranged from 19-52%, and the probability of 5 or more ranged from 13-71%, with long tails of outliers. However, with any interventions, this long tail of large outbreaks was substantially reduced.

**Figure 3:**
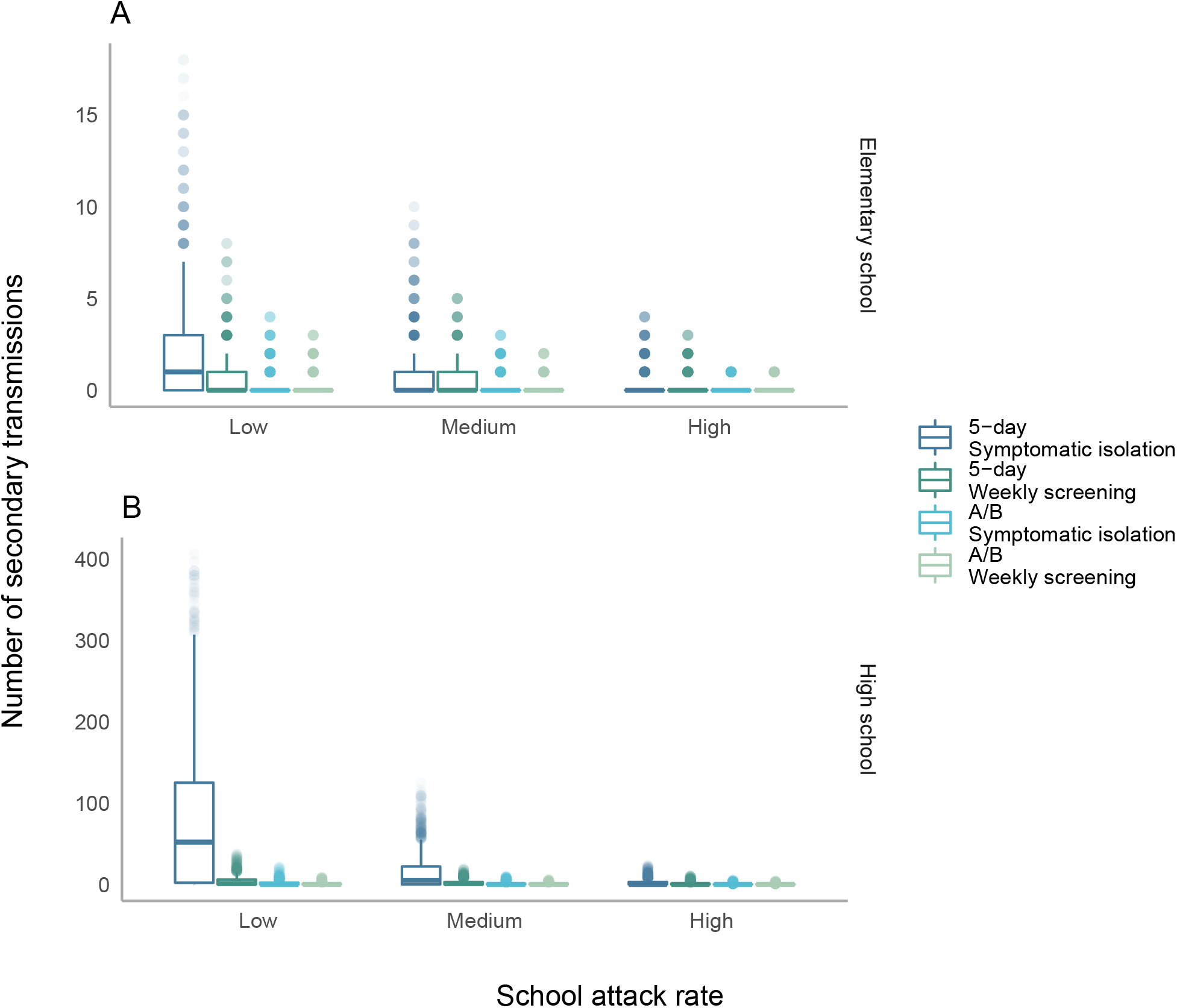
Distribution of secondary transmissions when a single case is introduced. The y-axis displays the number of secondary transmissions (outside of the index case’s household) when a case is introduced. Transmissions include both those directly from the index case, as well as those from secondary and tertiary cases. Distributions are truncated at the 99.5th quantile, i.e. all outcomes occur with at least probability 1/200.

### Transmissions over the course of the semester

With any of the modeled scenarios (symptomatic isolation, classroom quarantine, teacher vaccination, and weekly screening), both 5-day and A/B schedules increased transmission compared to remote learning. This increase was greater when community incidence was higher, especially considering infection risk among staff (Figures 4-5).

**Figure 4:**
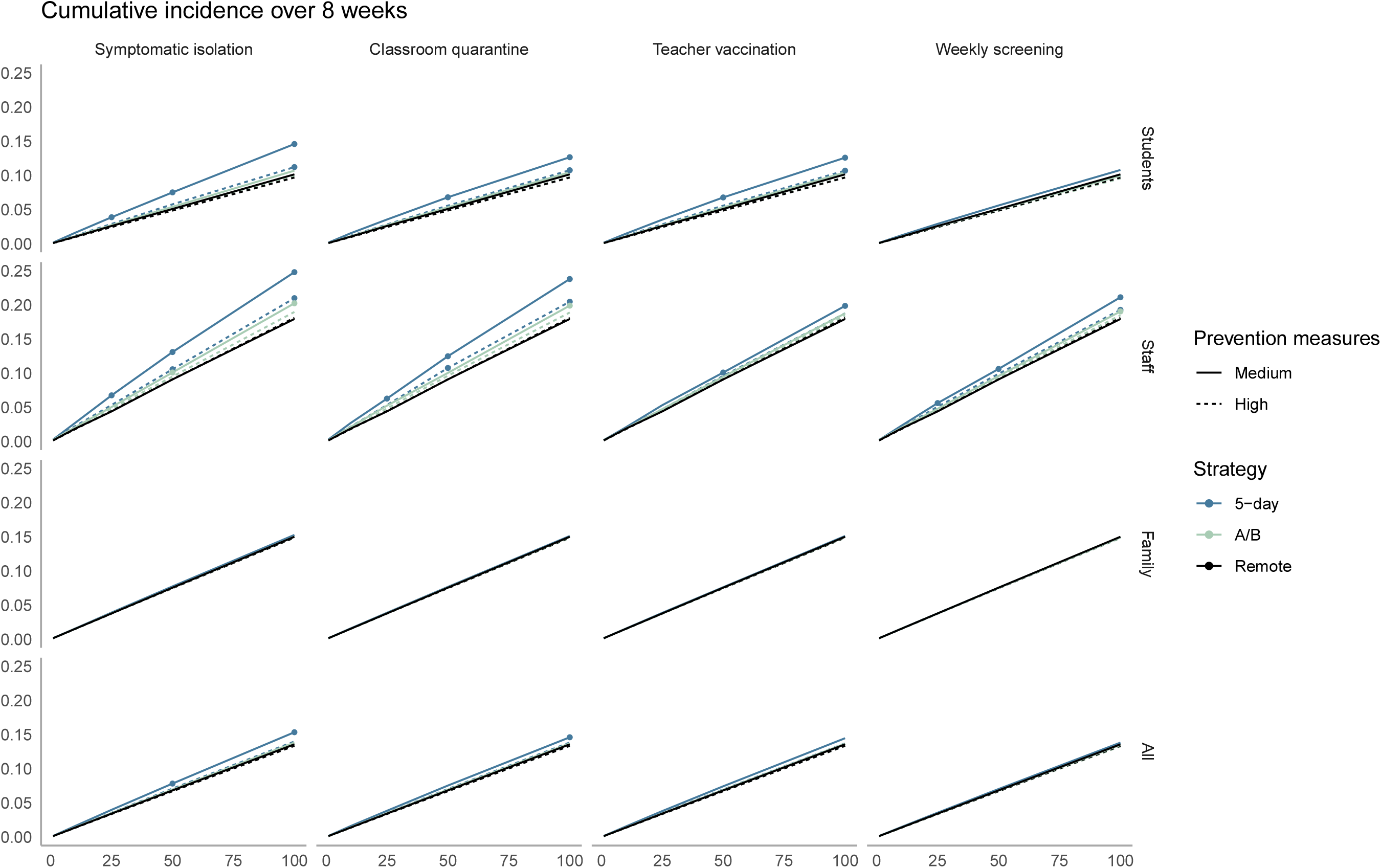
Cumulative incidence over 8 weeks in elementary schools. The x-axis shows the average daily community incidence per 100,000 population. The y-axis shows cumulative incidence over 8 weeks. Columns denote different isolation, quarantine, vaccination, and detection strategies, while rows show different population subgroups. Points are marked for strategies with increased incidence over remote learning that exceeds 1%.

Assuming that additional measures such as school closure were not adopted, Figures 4 and 5 display the values of community incidence at which our predefined threshold for “controlling transmission” was met. In elementary schools with medium mitigation under a 5-day schedule, all strategies met this threshold among when community incidence fell below 25 cases per 100,000 per day. The control threshold for the full population was exceeded at 50/100,000/day with symptomatic isolation (incidence 7.8%, increment 1.002%) and at 100/100,000/day with classroom quarantine (incidence 15%, increment 1.03% for classroom quarantine and 1.7% for symptomatic isolation). Evaluating the threshold specifically among teachers, the control threshold was never met when community incidence was *≥* 25 per 100,000/day, unless teachers were vaccinated (incidence: 6.2%, increment 1.9% with classroom quarantine). With both high mitigation and teacher vaccination, strategies met the control threshold among teachers at all rates up to 100 per 100,000 cases/day.

**Figure 5:**
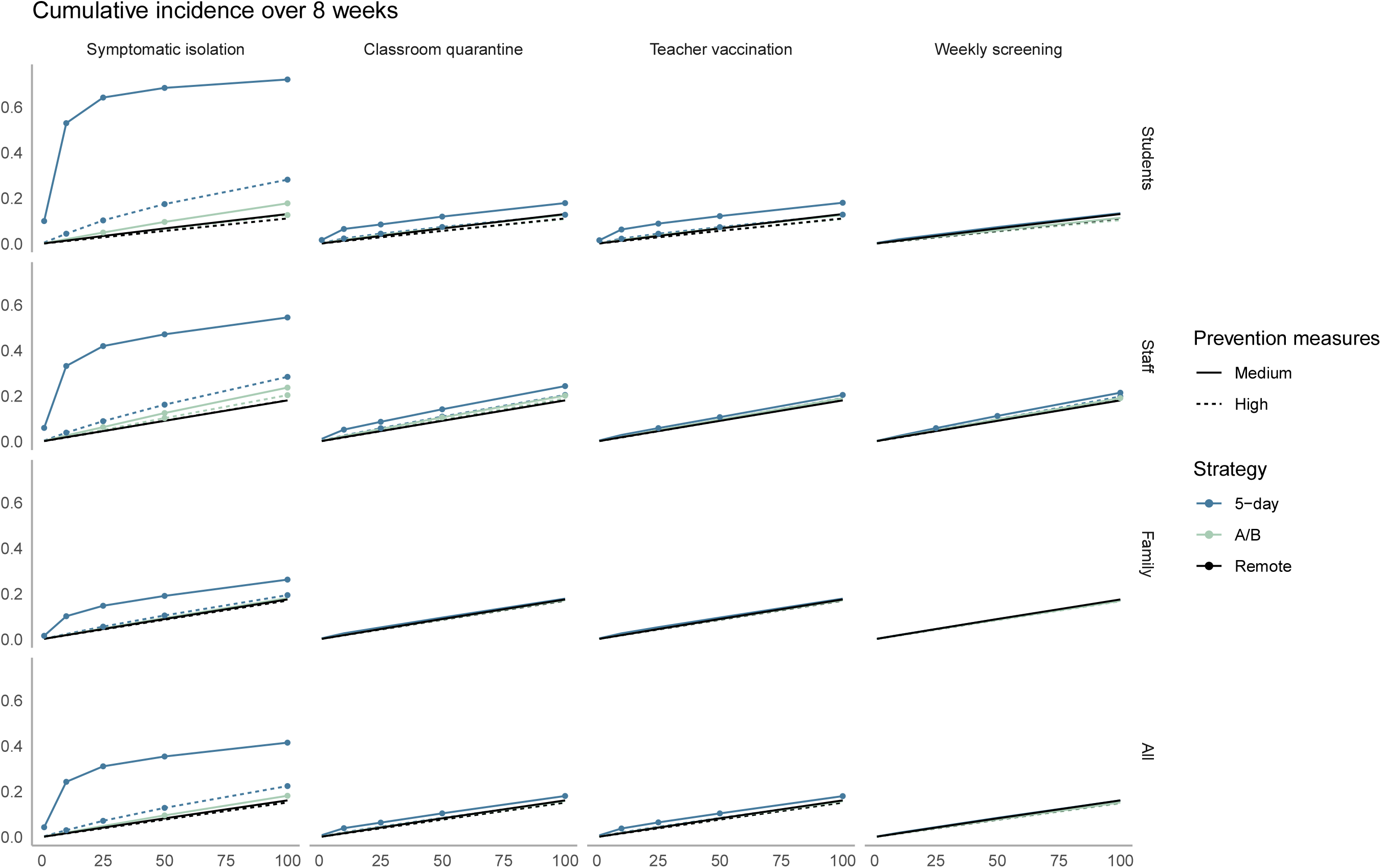
Cumulative incidence over 8 weeks in high schools. The x-axis shows the average daily community incidence per 100,000 population. The y-axis shows cumulative incidence over 8 weeks. Columns denote different isolation, quarantine, vaccination, and detection strategies, while rows show different population subgroups. Points are marked for strategies with increased incidence over remote learning that exceeds 1%.

In high schools, stronger mitigation or prevention strategies would be required to meet the control threshold. Under medium mitigation and community incidence of 100 cases per 100,000/day, only weekly screening met this threshold for the full population. Under high mitigation, all strategies except symptomatic isolation met this threshold. For the teacher control threshold in high schools, high mitigation combined with either weekly screening or teacher vaccination kept the increase in teacher cumulative incidence below the control threshold at 50 cases per 100,000/day.

### Sensitivity analyses

If children were as infectious as adults, the average number of secondary cases in elementary schools 30 days was 1.9 times higher compared to the base case; if adolescents were half as susceptible as adults, secondary cases in high schools were reduced by a factor of 0.3 (Figure S2, S3). When we repeated our analysis of elementary schools assuming equivalent variation in individual infectiousness for children as for adults, we found more instances of no onward transmission and a lower average number of secondary infections over 30 days, but also slightly larger outbreaks when they occurred (Figures S2, S3). After a single introduction, all types of 2-day schedules (e.g. MW/ThF vs. MT/WTh) led to similar numbers of secondary infections, similar chance of any secondary infection, and similar numbers of secondary infections among teachers/staff (Figures S2, S2). The benefits of hybrid scheduling generally persisted across increased levels of out-of-school mixing (Figures S5-S6). In order for student and caregiver secondary infections to be greater with an A/B model than a 5-day model, >9 families needed to interact on each day out of school.

## DISCUSSION

While in-person education poses some COVID-19 transmission risk, the results of this simulation model underscore that it is possible to offset this with adequate precautions, particularly in elementary schools and when community transmission is well-controlled. In elementary schools with adherence to masking and distancing, our results demonstrate most cases introduced into a school would lead to little or no onward transmission. Nevertheless, if transmission occurs, it may be difficult to link to the school, and even modeled scenarios that commonly lead to no in-school transmission can occasionally generate larger outbreaks in a school community. As other models have noted (28,30), the risk of this is substantially higher in high schools compared to elementary schools.

While it is difficult to determine the exact risk of these low-probability but high-consequence events, community transmission determines the number of cases that enter a school community and therefore such rare but consequential outbreaks become increasingly likely when local incidence is high. The guidance from the US Centers for Disease Control and Prevention regarding K-12 education highlights population-adjusted incidence as a core indicator (50), and our results provide additional evidence supporting use of this metric for school decisions in the absence of in-school screening or surveillance data. Nevertheless, while schools may decide that in-school transmission risk is too high above certain levels of community incidence, our results do not suggest that K-12 education with mitigation and/or modified schedules, is likely to be a primary driver of sustained community transmission. In the event that community-level R(t) exceeds 1, it may be possible to lower this through targeted closure of high risk venues, and maintain R(t) below 1 while schools remain open.

Our results are compatible with global observations regarding school outbreaks, in which numerous well-studied index cases have produced no or minimal secondary cases, but larger outbreaks have also been recorded, particularly in secondary schools (10,51,52). Teachers and staff tend to be over-represented in school outbreaks relative to their presence in a school community. At the elementary level, they often represent a third or more of diagnosed secondary cases (7,12). As a substantial fraction of staff may be at high risk of complications, it is important that schools undertake precautions specifically to prevent transmission among staff. These may include holding faculty meetings virtually, mandating masks during all work in shared spaces, and establishing locations for lunches and mask breaks that permit distance and ventilation. Hospitals have similarly found patterns of staff-to-staff transmission and implemented measures such as reducing opportunities for communal food consumption (53). It is also critical that staff have adequate financial and logistical support to isolate when exhibiting symptoms and to quarantine following COVID-19 exposure, even if exposed outside of the school setting.

We predict that most in-school transmission will occur in the classroom during sustained contact, and consequently interventions which reduce classroom transmission can be highly effective, such as distancing, masking, or reducing class sizes. The latter may be achieved through the addition of more staff and moving into previously unused spaces, by allowing families to opt out of in-person learning, by prioritizing a subset of vulnerable students for limited in-person slots, or through implementation of hybrid scheduling. Some have raised concerns that a hybrid model could paradoxically increase SARS-CoV-2 transmission in schools by leading to greater out-of-cohort mixing on days when children are not physically in school. However, we find that the A/B schedule leads to far fewer infections in the school community than a 5-day schedule, a result that persists even with an assumption of substantial out-of-school mixing between families. Weekly SARS-CoV-2 testing of asymptomatic people (screening) could offer benefits comparable to an A/B schedule, and thereby allow more in-person instruction with less disruption.

Weekly asymptomatic SARS-CoV-2 screening is particularly valuable in light of uncertainty in model parameters and outcomes. There is still debate surrounding relative susceptibility and transmissibility of children compared to adults. In addition, the degree of variation in individual-level infectiousness is uncertain. Such variation can arise from biologic factors associated with viral control, perhaps ACE2 receptor density, or from biologic or behavioral factors that lead to spread of respiratory droplets such as body size, cough strength, and tendency to contain coughs or sneezes (48,54–57). Increases in this “overdispersion” lead to a wider range of possible outcomes when an infection is introduced, including both greater instances of zero in-school transmission as well as larger outbreaks when secondary transmission occurs. This variability in possible outcomes is markedly reduced when hybrid schedules and/or weekly screening are implemented.

Beyond biological uncertainty, it may also be difficult to ascertain local behavioral parameters such as adherence to masking, distancing, and quarantine protocols, which can vary across different settings and over time. Coupled with stochastic uncertainty in outbreak size and a high proportion of subclinical illnesses in school populations, it may be difficult to confidently ensure minimal transmission and detect outbreaks that occur before they spread to the community. In particular, the low probability of clinical disease for infected children means that transmission chains in schools are likely to be only partially observed, and linked cases may be mistakenly classified as isolated introductions. While regular screening can both improve these data and prevent transmission through early detection, at a minimum, schools should be on alert for signs that an outbreak is brewing undetected, and consider screening of all students and staff in response to the detection of any case without a clearly identified in-school or out-of-school link. Other factors not considered in this work that may affect transmission include changes in parent behavior when children return to school and substitution effects in staff contacts when school is remote or hybrid.

Last, a new variant of COVID-19 (B.1.1.7) has recently been identified that is approximately 50% more transmissible than the strains that have previously dominated, with possibly larger increases in transmissibility among children than among adults compared to the previous strain (58). This variant has been linked to a large outbreak in a primary school in the Netherlands and triggered a new wave of school closures in Europe (19,59). If this or other similar variants spread throughout the United States, classroom-based efforts at infection prevention such as masking and distancing may be less effective at suppressing attack rates. For instance, schools which have currently achieved attack rates commensurate with “medium uptake of mitigation” may find that the same measures will result in attack rates reflecting “low uptake.” This shift points to the even more urgent need for data and underscores the added value of routine asymptomatic screening.

There are inevitable trade-offs between school disruption, risk of in-school transmission, and resources required to implement in-school prevention approaches. Decisions about the acceptable balance between these trade-offs are best made by school communities. We offer a quantitative perspective on the way that in-school transmissions are likely to occur, as well as the impact that proposed protocols could have on that transmission risk. We emphasize that particularly among young children, schools appear to be ‘mirrors’ of community transmission, rather than ‘amplifiers’ or ‘brakes.’ Thus, a reliable way to ensure a low infection risk in school is to reduce infectious introductions, by keeping community incidence low (28). However, even when introductions occur, high adherence to in-school prevention measures, preferably complemented by regular asymptomatic testing and teacher vaccination, can also permit return to in-person education with controlled risk of in-school COVID-19 transmission. Local, state, and federal agencies should prioritize these effective interventions that permit the benefits of in-person education while protecting the safety of both students and educators.

## Data Availability

Model code is available at https://github.com/abilinski/BackToSchool2.

## Supplement

**Figure S1:**
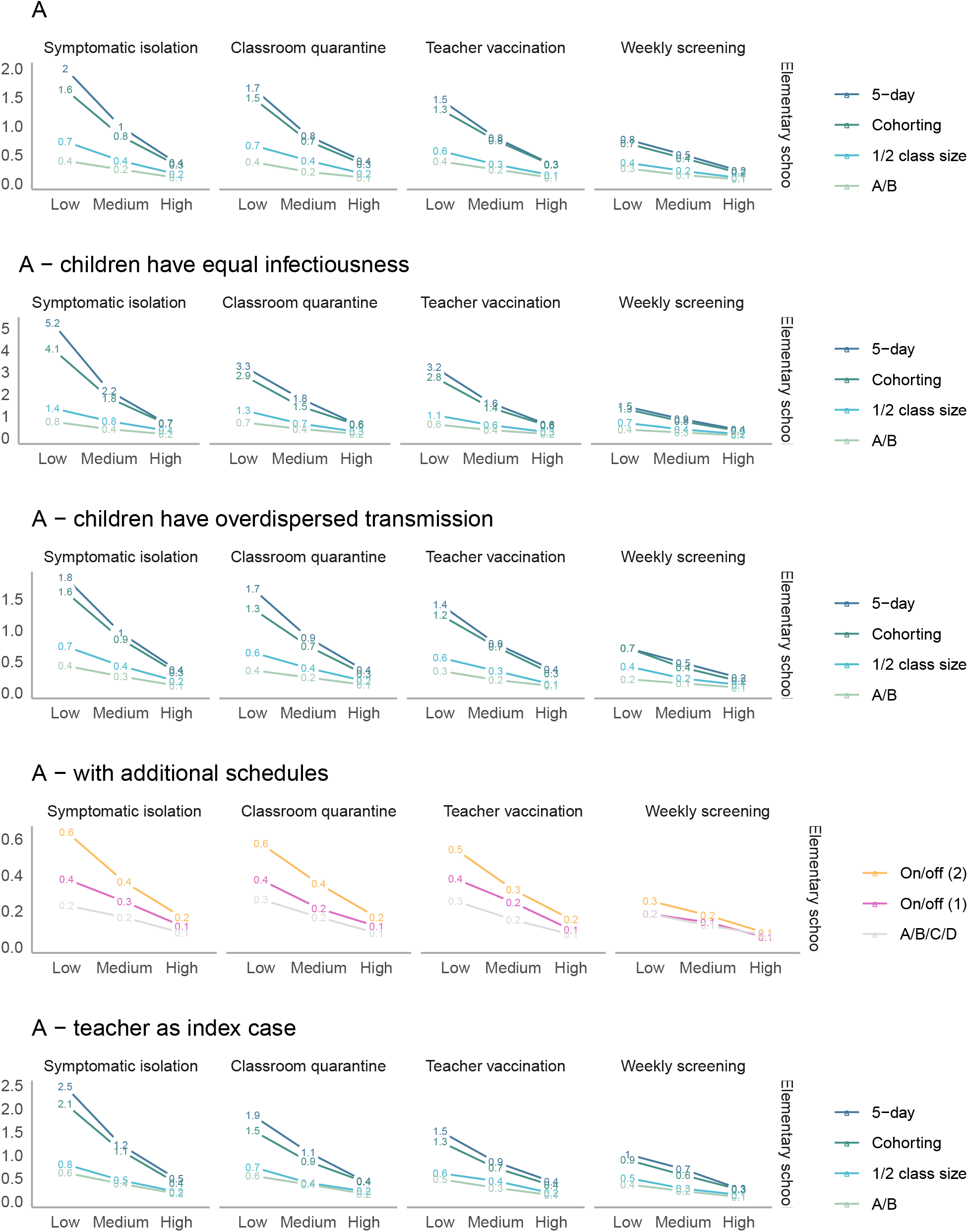
Sensitivity analyses (elementary schools) – average number of total secondary transmissions over 30 days (outside of the index case’s household) following a single introduction into a school community. The columns vary the level of prevention measure uptake, with low uptake assuming minimal interventions and high uptake assuming intensive interventions. Line colors correspond to scheduling strategies.

**Figure S2:**
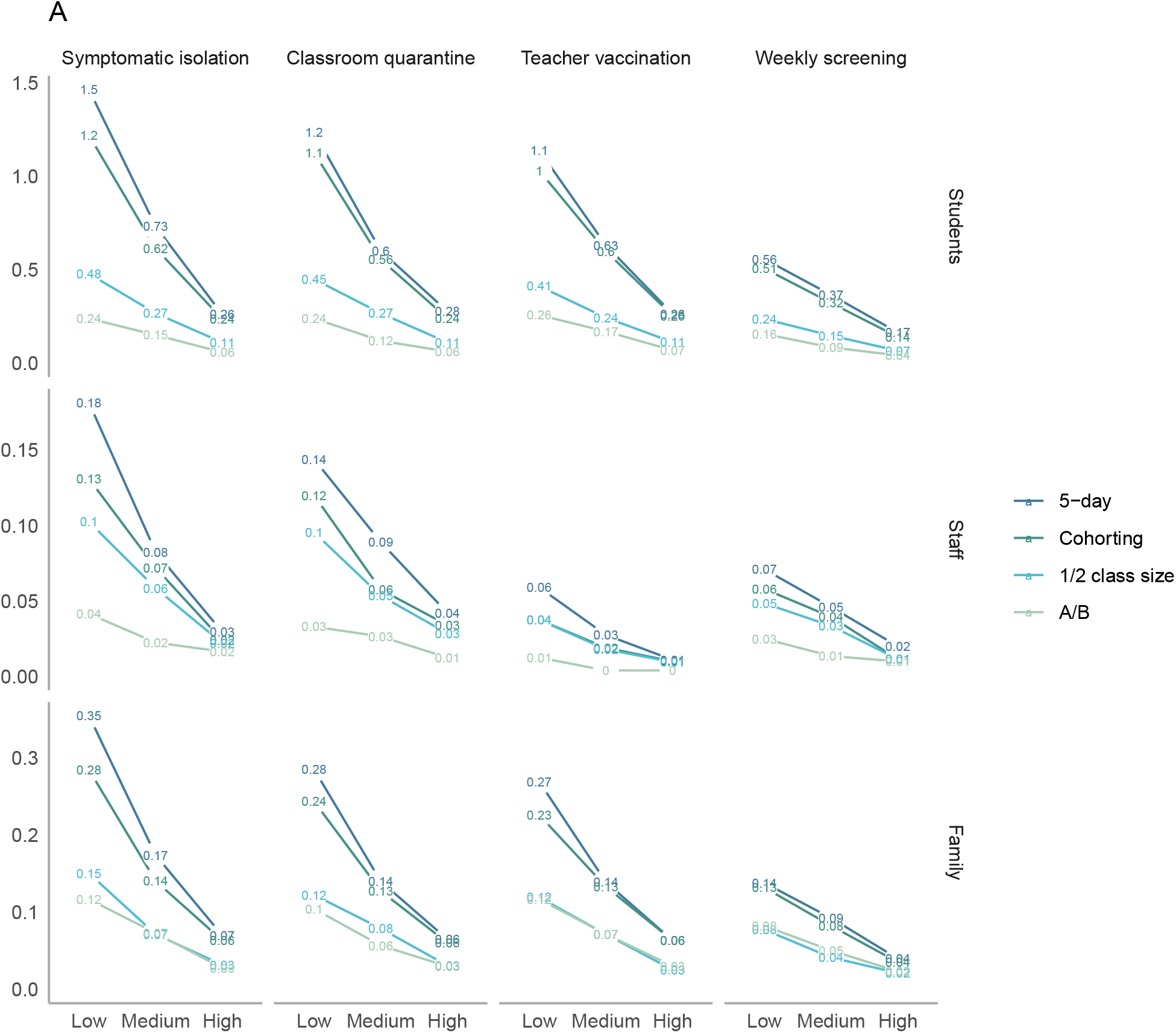
Sensitivity analysis - elementary schools by case type. Average number of total secondary transmissions over 30 days (outside of the index case’s household) following a single introduction into an elementary school community. These include both transmission directly from the index case, as well as from secondary and tertiary cases. The x-axes varies the level of prevention measure uptake, with low uptake assuming minimal interventions and high uptake assuming intensive interventions. Line colors correspond to scheduling strategies.

**Figure S3:**
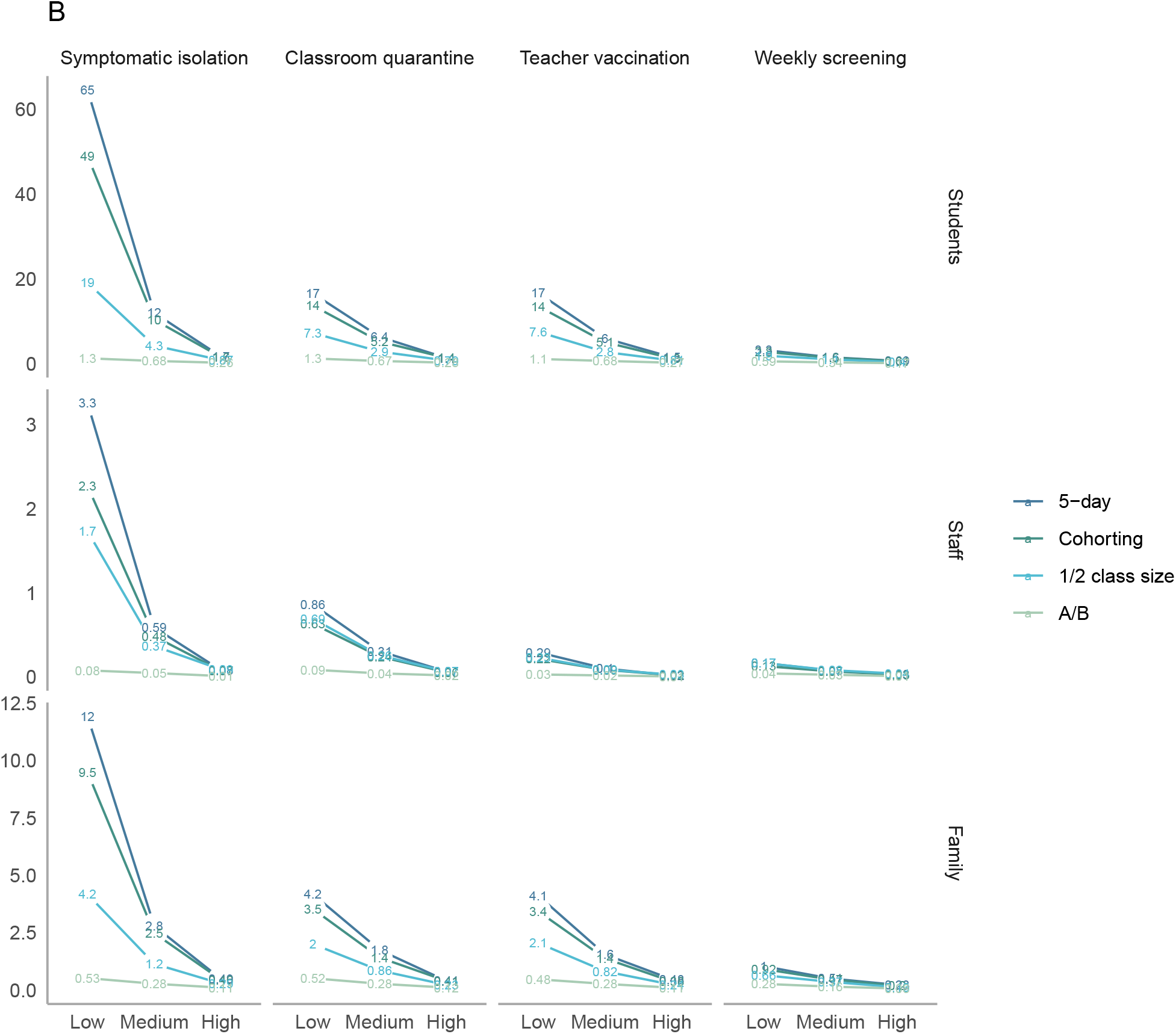
Sensitivity analysis - high schools by case type. Average number of total secondary transmissions over 30 days (outside of the index case’s household) following a single introduction into an elementary school community. These include both transmission directly from the index case, as well as from secondary and tertiary cases. The x-axes varies the level of prevention measure uptake, with low uptake assuming minimal interventions and high uptake assuming intensive interventions. Line colors correspond to scheduling strategies.

**Figure S4:**
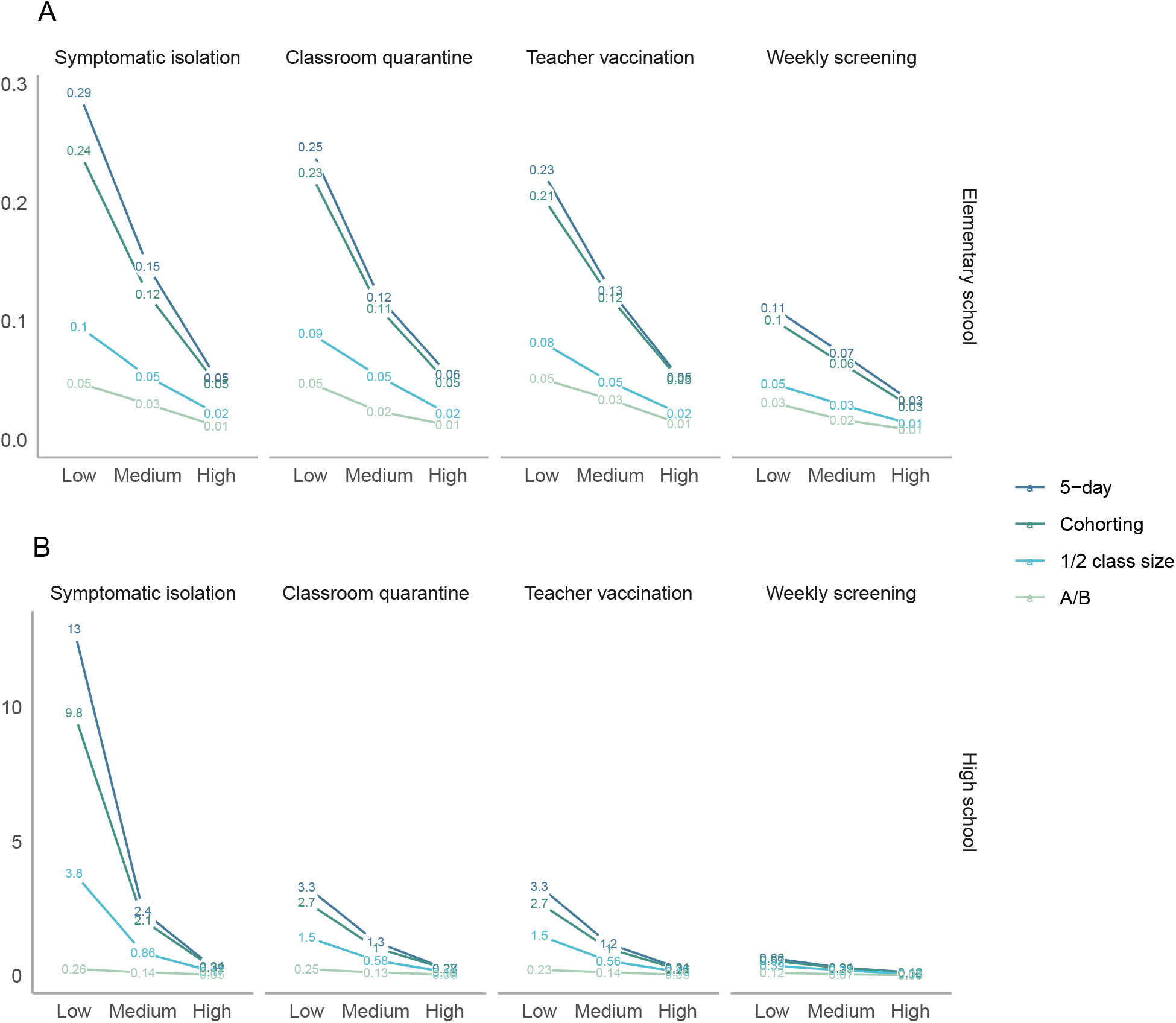
Average number of clinically symptomatic cases in staff and students over 30 days following a single introduction into a school community. These include both transmission directly from the index case, as well as from secondary and tertiary cases. The top panel shows elementary schools, where children are assumed to be less susceptible and less infectious, while the bottom panel shows high schools. Note that axes differ across rows. The x-axes vary the level of prevention measure uptake, with low uptake assuming minimal interventions and high uptake assuming intensive interventions. Line colors correspond to scheduling strategies.

**Figure S5:**
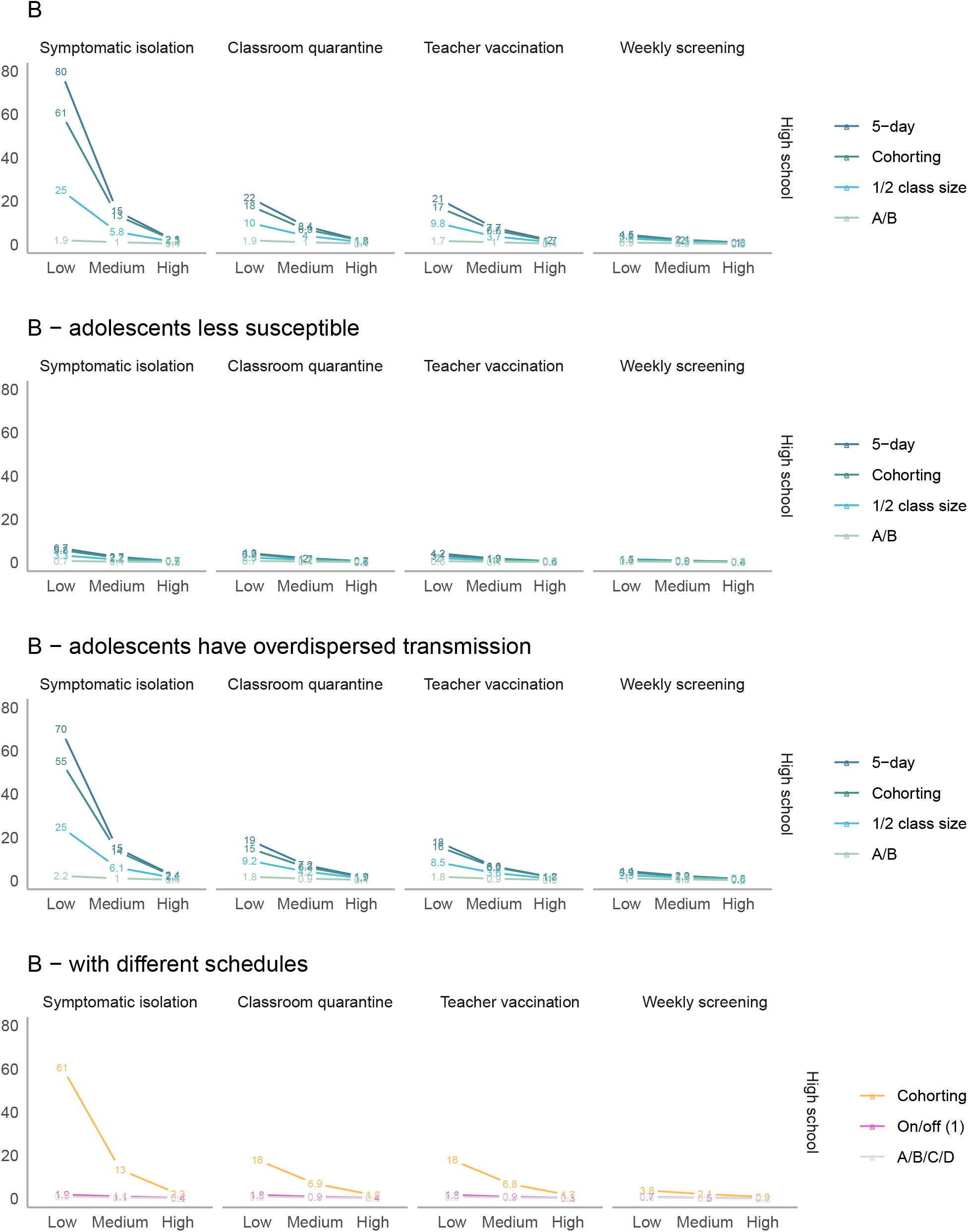
Sensitivity analyses (high schools) – average number of total secondary transmissions over 30 days (outside of the index case’s household) following a single introduction into a school community. The columns vary the level of prevention measure uptake, with low uptake assuming minimal interventions and high uptake assuming intensive interventions. Line colors correspond to scheduling strategies.

**Figure S6:**
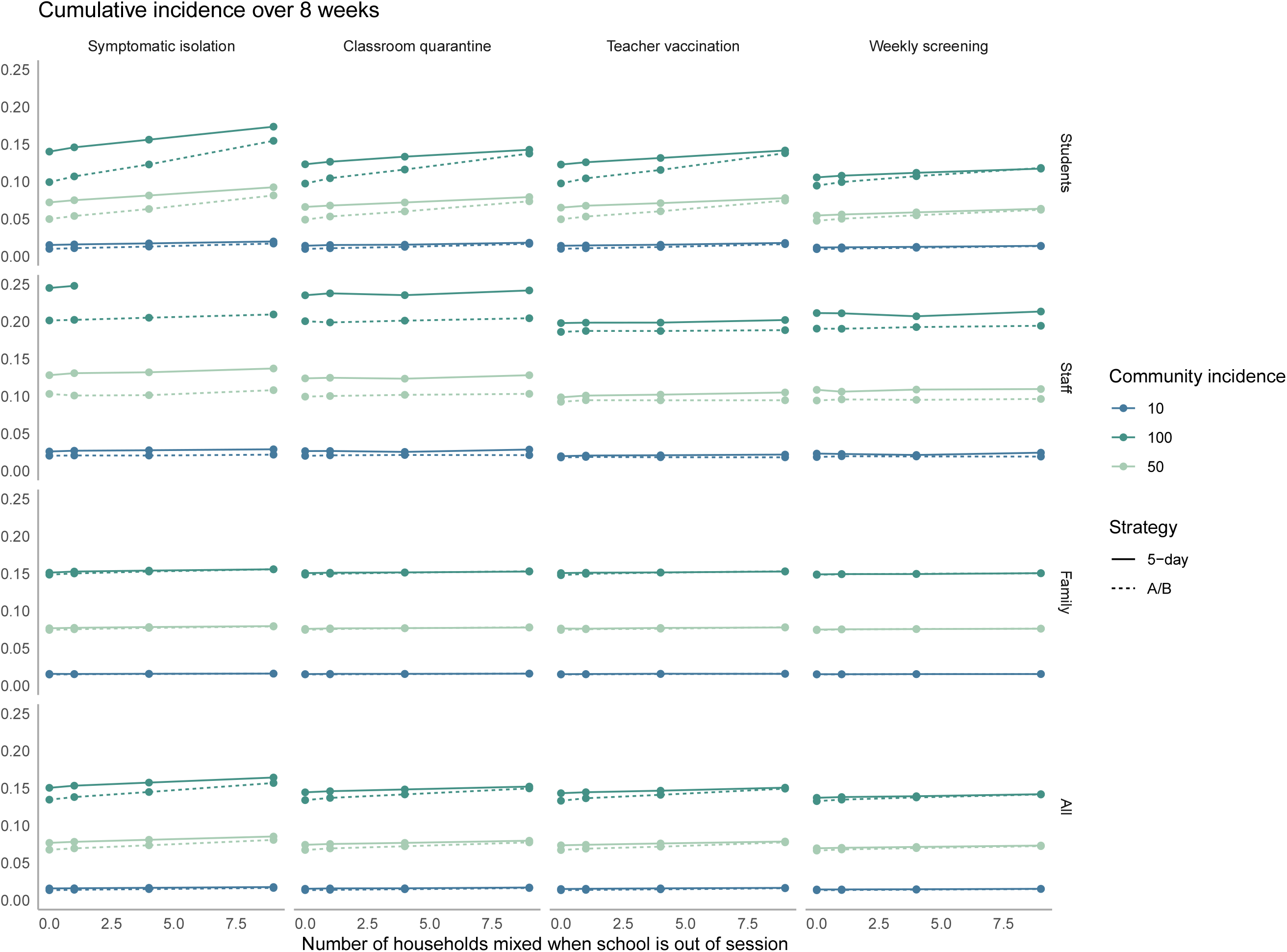
Cumulative incidence over 8 weeks in elementary schools across different levels of out-of-school mixing. The x-axis shows the average daily community incidence per 100,000 population. The y-axis shows cumulative incidence over 8 weeks. Columns denote different isolation, quarantine, vaccination, and detection strategies, while rows show different population subgroups

**Figure S7:**
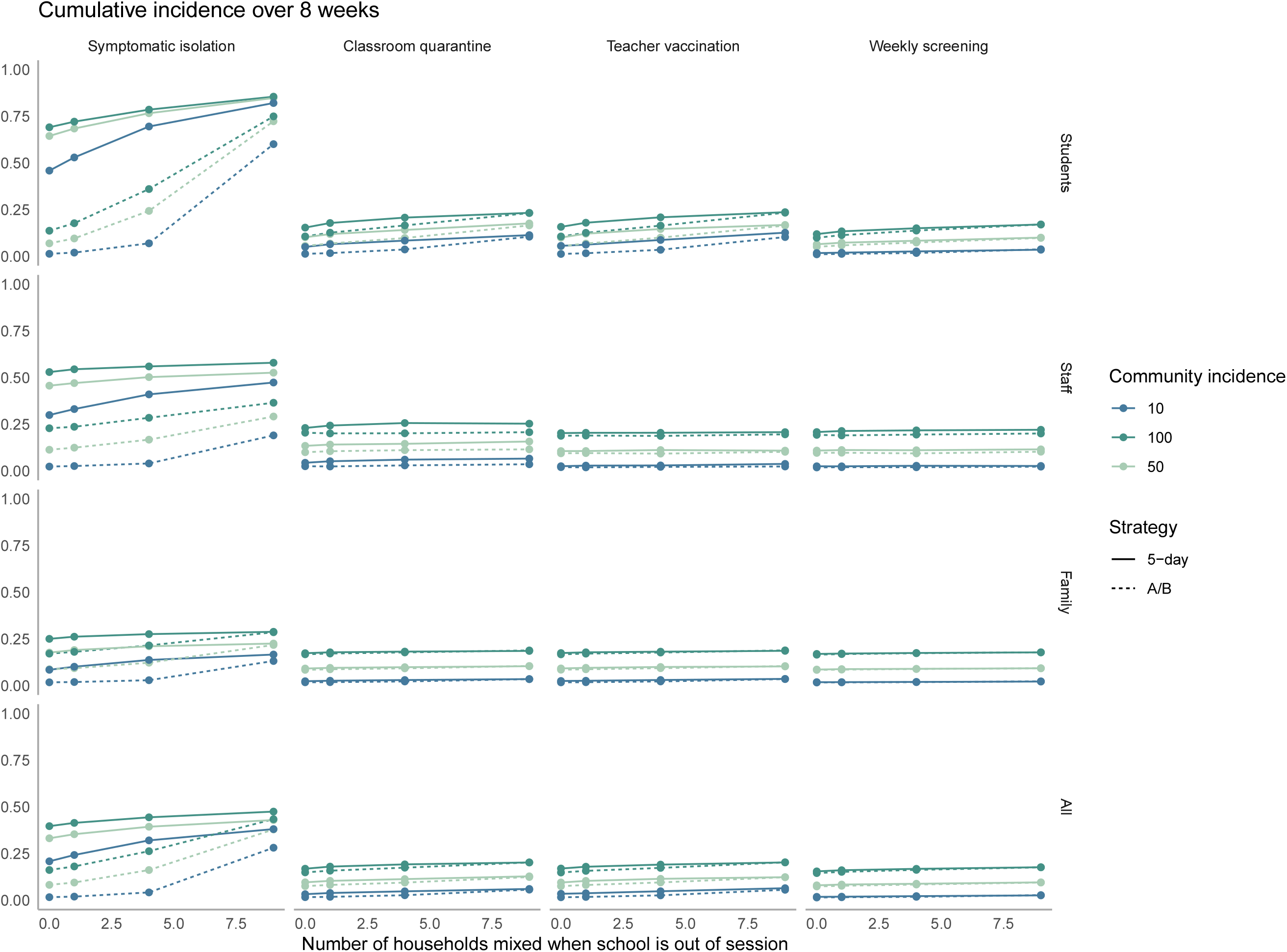
Cumulative incidence over 8 weeks in high schools across different levels of out-of-school mixing. The x-axis shows the average daily community incidence per 100,000 population. The y-axis shows cumulative incidence over 8 weeks. Columns denote different isolation, quarantine, vaccination, and detection strategies, while rows show different population subgroups.

## Model

We use a Framework for Reconstructing Epidemiological Dynamics (FRED) to generate household structures (36). For computational simplicity, we used Maryland as a representative state, as sibling structure (the main parameter of interest) did not appear sensitive to location.

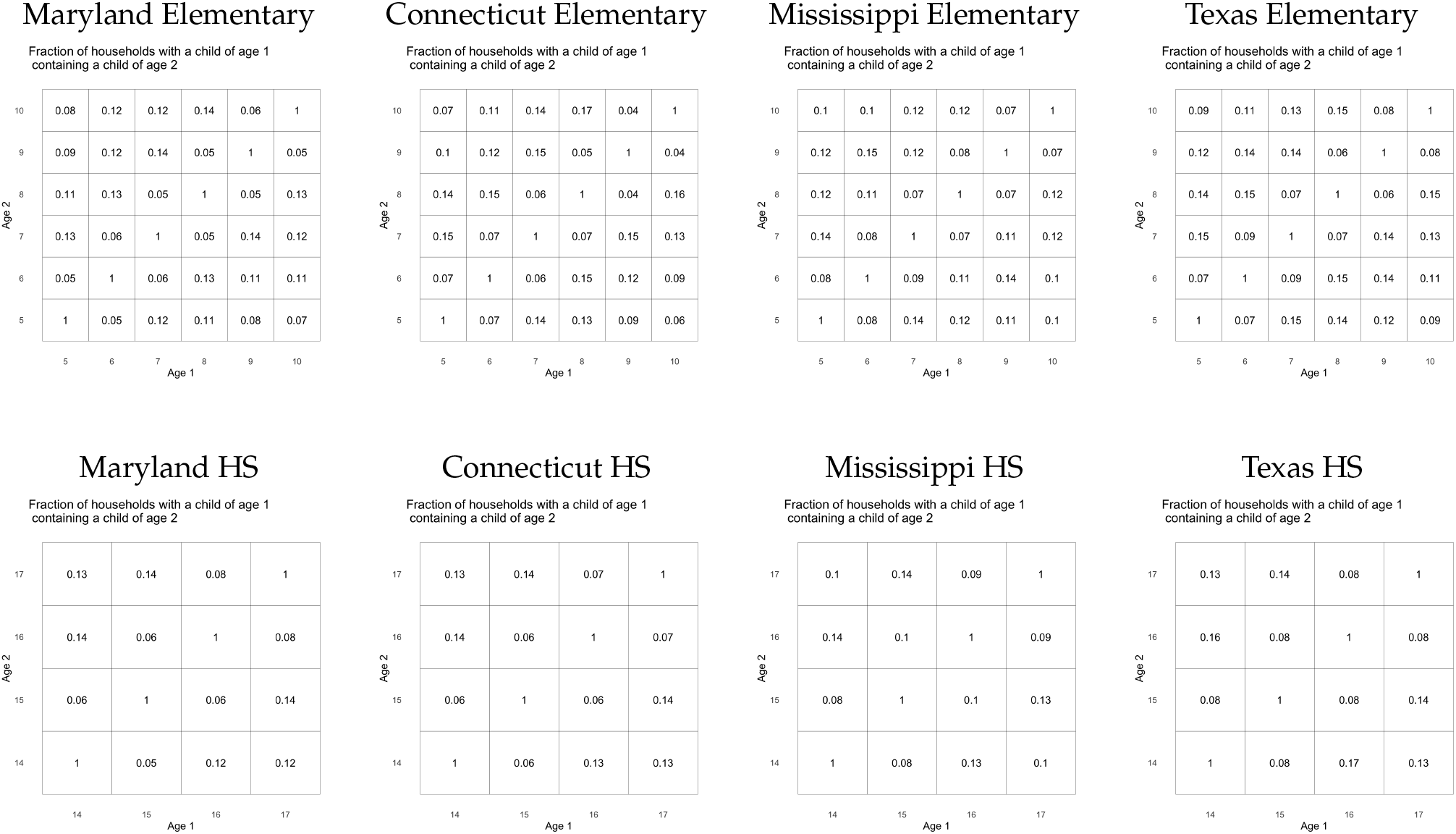

### Comparison to observed outbreaks

A number of factors make formal calibration challenging for this paper. First, most data collection has been ad hoc, with some sources biased toward reporting large outbreaks and others toward high-mitigation schools who voluntarily collect and report data. Without comprehensive data on school mitigation efforts, interpretation can be challenging, as our results emphasize that a large range of outcomes is possible due to both variation in parameters and stochastic uncertainty. Testing practices vary across schools, affecting reporting procedures, and the definition of “contact” also varies, with some sources including even brief contacts while others require more sustained interaction (e.g. >15 minutes without a mask). Overall, this section describes available data sources in comparison to our results. We emphasize that substantial uncertainty persists, and that weekly screening or other surveillance is one of the best tools available for understanding a specific context as well as early detection of outbreaks.

### Differences in infectiousness and susceptibility by age

In our model, we assumed that young children (10 and under) were both less susceptible and less infectious than adults. To inform these assumptions, we used a meta-analysis on child susceptibility on those under 18-20 (43), which was consistent with best-fit model estimates (67) and another on child infectiousness (68). We also used a number of contact tracing studies suggesting not just a difference between children and adults but also an age gradient in susceptibility and infectiousness. These included a study from B’nei Brak, Israel on household infections (Figure 4) (44) and two studies from France contrasting minimal elementary school outbreaks (despite introductions) with a larger high school cluster in an area with early COVID-19 exposure (8,45). Limited data from Iceland, with comprehensive contact tracing and sequencing, suggested a similar difference between young children and adolescents in infectiousness (69). Contact tracing data from South Korea on infectiousness, while more difficult to interpret due to concurrent exposures among household members and high PPE usage of guardians of infected children (70,71), was not inconsistent with this finding (31).

We focused on data from contact tracing studies that used comprehensive testing of contacts because these were less likely to be biased. In particular, we did not want to interpret evidence that children were rarely identified as index cases (68) or had lower seroprevalence in some contexts as evidence that they were less susceptible or infectious (72). These differences could have been driven by the fact that children are less likely to have symptoms and be tested and/or that their contacts were markedly reduced by school closures early in the pandemic. While household contact tracing studies with comprehensive testing avoided this issue, they could have had other biases. For example, children were unlikely to be caretakers of sick individuals commpared to adults in the household, and may have been shielded in houses with known cases, particularly in the midst of an unprecedented pandemic. Nevertheless, in general, higher household attack rates in children have been observed for seasonal influenza and H1N1, making lower estimates for COVID-19 particularly notable (73–78).

These findings nevertheless cannot differentiate between biological explanations for lower susceptibility in younger children (e.g., lower density of ACE2 receptor) and behavioral ones (e.g., easier to restrict socialization). In addition, some studies suggest higher susceptibility and/or infectiousness of young children than we include in our base case (79–83). While we model these possibilities in sensitivity analyses, the bulk of evidence on well-studied school outbreaks has pointed to important distinctions between elementary and high school-aged children. For example, when Israel experienced significant outbreaks upon return to school in the early summer, there was a significant outbreak of 178 cases in a middle/high school (concentrated in grades 7-9), but elementary school outbreaks were generally reported as smaller (e.g. 33 cases) (10,18). This difference was also apparent in informal databases, with high schools largely responsible for outbreaks of more than 50 people (e.g. in New Zealand and the US prior to social distancing and Australia, where a school outbreak was reported to be driven by high schoolers) (84–86). In the Netherlands, a health official was quoted saying that significant outbreaks occurred mainly in high schools and universities prior to an elementary school outbreak with B.1.1.7 (59). Exceptions often included significant outbreaks among teachers (e.g., in Chile (12) and Singapore (87)).

### Secondary cases

Our results are broadly consistent a few key features of observed data. First, well-studied cases have led to no or minimal outbreaks in a number of settings. In passive surveillance from the United Kindom during the summer, in-school transmission was identified from 39% of index cases in secondary schools and 26% of cases in primary schools “in the context of small class or bubble sizes, half empty schools, and extensive hygiene measures.” This is similar to what we predicted with an A/B model in secondary schools with medium mitigation (36%) and a low mitigation scenario in elementary schools (23%). No onward transmission was found in Singapore or Ireland, each with 3 seed cases (9,88). In Rhode Island childcare settings, which had small class sizes, onward transmission was documented in 4/29 index cases (14%), consistent with 1/2 class size scenario and high mitigation (12%) (51). However limited testing in some of these studies may have missed subclinical cases.

Second, several data sources showed signs of overdispersion with the possibility of large outbreaks alongside cases without apparent transmission. In the Rhode Island example, one outbreak involved 10 cases among contacts (10 children, four staff members, and one parent); in another study from Australia, 9 cases in early childhood education centers led to no onward transmission while one led to 13 infections (7). If these were indeed caused by a single index case, the level of overdispersion in our model may not capture such extreme outcomes.

Last, teachers were often overrepresented in outbreaks even in well-studied outbreaks, with 16% of staff and 10% of students having antibodies in a Chilean outbreak across multiple school levels (12). In the Australian study, adults comprised 8/18 of secondary cases identified (7).

### Frequency of infections and subclinical infections in children compared to adults

Our model predicted both lower incidence of infections in children and a higher rate of underdiagnosis. With these combined, we would expect to see fewer cases in children, but a smaller relative difference when comprehensive surveillance and/or random testing is conducted, which is consistent with observed data. For example, in passive surveillance from the United Kingdom, staff had more than 4 times the COVID incidence of students per 100,000 across all age groups; however in random testing, observed prevalence was roughly equal among students and staff (89,90). In elementary schools in New York in fall 2020, schools reported substantially fewer cases in elementary school students than in staff per population, but in random surveillance testing from the same period in New York City (manually extracted and analyzed by others), prevalence was roughly equal in students and staff (91,92). While these are difficult to compare directly as people who self-isolate for symptoms are not present for random testing, the contrast remains striking. Less systematically, a major outbreak in an Israeli high school was detected from wide-scale testing after observing 2 unlinked community cases (10), and the first Ontario school to participate in voluntary mass asymptomatic screening closed after uncovering a substantial number of previously undetected cases (93).

### Effective reproduction number

In Figure S8, we display the effective reproduction number associated with different scenarios. One modeling study estimated that from August through October 2020, there was an average effective reproduction number of 0.54 [0.44-0.62] for children 0-9 and 0.75 [0.59-0.89] for children 10-19 (94). As noted in the introduction of this article, school openings varied considerably across the country, but both of these fall within our range of estimates. For elementary schools, it is consistent with full opening and high mitigation, hybrid opening or limited attendance and medium mitigation, or some combination of these with remote school. For high schools, it is most consistent with high mitigation limited attendance or a hybrid model or some combination of these with remote school.

**Figure S8:**
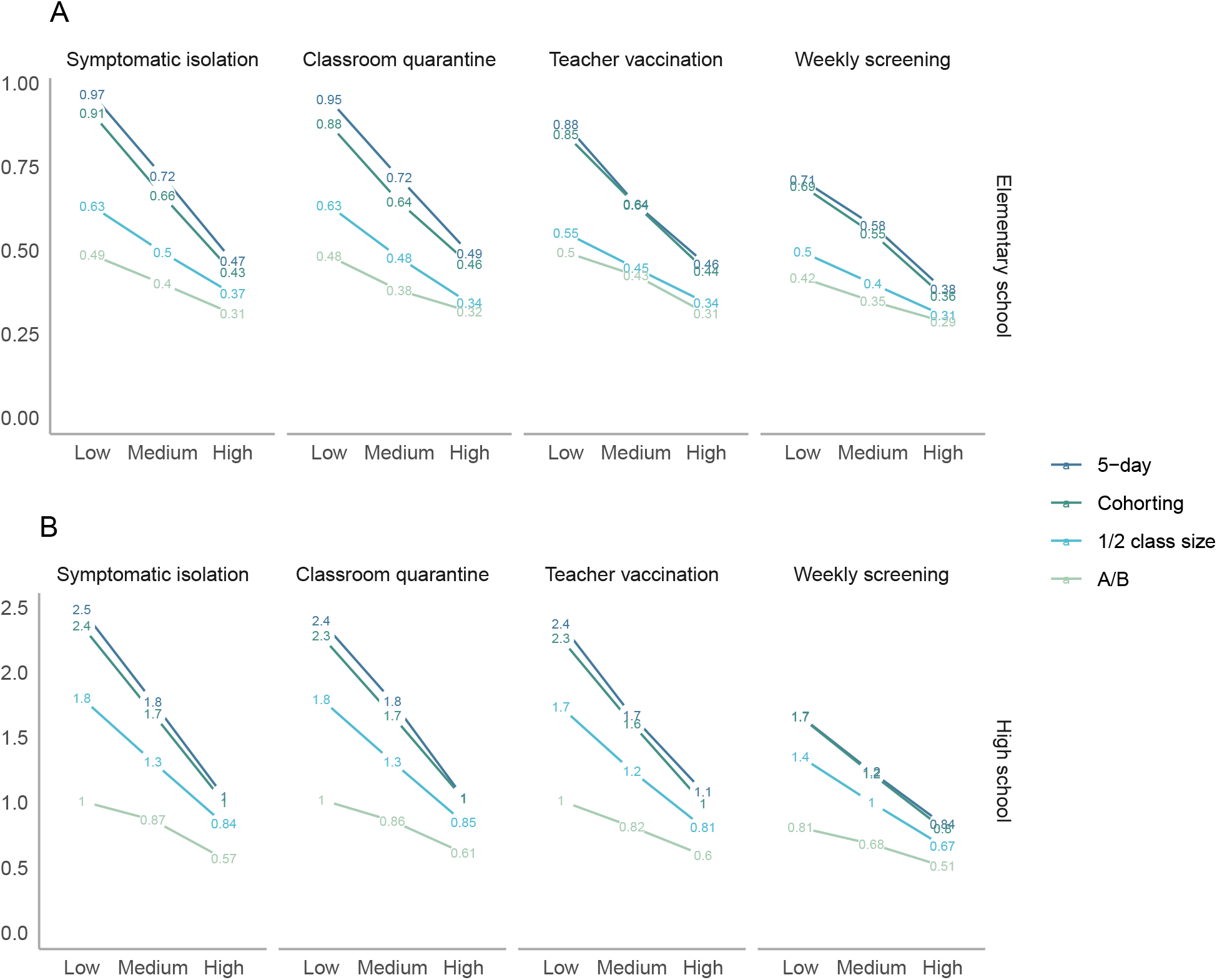
Effective reproduction number, defined as average secondary transmissions following a single introduction into the school community. The top panel shows elementary schools, where children are assumed to be less susceptible and less infectious, while the bottom panel shows high schools. Note that axes differ across rows. The x-axes vary the level of prevention measure uptake, with low uptake assuming minimal interventions and high uptake assuming intensive interventions. Line colors correspond to scheduling strategies.

### Population-level studies

While we do not directly model full community incidence, two recent studies using quasi-experimental data to study the impact of school reopening on transmission were consistent with our observation that there is a higher risk of increased community transmission following school reopenings when initial transmission was high, contrasted to tight null effects when initial transmission was lower (95,96).

## Acknowledgements

The authors were supported by the Centers for Disease Control and Prevention though the Council of State and Territorial Epidemiologists (NU38OT000297-02; AB, JAS), the National Institute of Allergy and Infectious Diseases (R37AI058736-16S1; ALC and K01AI141576; MCF), the National Institute on Drug Abuse (3R37DA01561217S1; JAS). The papers’ contents are solely the responsibility of the authors and do not necessarily represent the official views of the funders. Model code is available at https://github.com/abilinski/BackToSchool2.

## Footnotes

5 When applied to a 5-day school week, the average adult-adult attack rate over the full duration of infection was 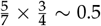 that of the household attack rate.

